# T_1_/T_2_ Ratio Imaging Improves Cortical Lesion Contrast in Multiple Sclerosis on 3T MRI

**DOI:** 10.1101/2022.03.24.22272914

**Authors:** Abigail R Manning, Erin S. Beck, Matthew K. Schindler, Govind Nair, Prasanna Parvathaneni, Daniel S. Reich, Russell T. Shinohara, Andrew J. Solomon

## Abstract

**Background:** Cortical demyelinated lesions are prevalent in multiple sclerosis (MS) and associated with disability; their presence on MRI has recently been incorporated into MS diagnostic criteria. Presently, advanced and ultra-high-field MRI — not routinely available in clinical practice — are the most sensitive methods for detection of cortical lesions, and approaches utilizing MRI sequences obtainable in routine clinical practice remain an unmet need.

**Objective:** To assess the sensitivity of the ratio of T_1_-weighted and T_2_-weighted (T_1_/T_2_) signal intensity for focal cortical lesions in comparison to other established, sensitive, advanced and high-field imaging methods.

**Methods:** 3-tesla (3T) and 7-tesla (7T) MRI collected from 10 adults with MS participating in a natural history study at the National Institutes of Health were included in the study. T_1_/T_2_ images were calculated by dividing 3T T_1_w images by 3T T_2_w fluid-attenuated inversion recovery (FLAIR) images for each participant. Cortical lesions were identified using 7T T_2_*w and T_1_w images and corresponding voxels were assessed on registered 3T images. For each participant, ratios derived from the median signal intensity of nonlesional tissue in the cortical region of the lesion and the median lesional voxel intensity were computed. These values were compared across 3T imaging sequences, including the calculated T_1_/T_2_ image, as well as T_1_w, T_2_w, and Inversion Recovery Susceptibility Weighted Imaging with Enhanced T_2_ weighting (IR-SWIET) images.

**Results:** 614 cortical lesions were identified on 7T images. 3T T_1_/T_2_ images demonstrated a larger contrast between median nonlesional cortical signal intensity and median cortical lesion signal intensity (median ratio = 1.29, range 1.19 – 1.38) when compared to T_1_w (1.01, 0.97 – 1.10, p<0.002), T_2_w (1.17, 1.07 – 1.26, p<0.002), and IR-SWIET (1.21, 1.01 – 1.29, p<0.03).

**Conclusion:** T_1_/T_2_ images are sensitive to cortical lesions. Approaches incorporating T_1_/T_2_ could improve the accessibility of cortical lesion detection in research settings and clinical practice.

## INTRODUCTION

Multiple sclerosis (MS) is a chronic inflammatory disease characterized by demyelinating lesions of the central nervous system. Cortical lesions in MS are an important contributor associated with disability^1-4^. The identification of cortical lesions was also incorporated into the 2017 revisions to MS diagnostic criteria^5^. However, MRI sequences routinely obtained in clinical care are largely insensitive to cortical lesions, especially intracortical and subpial lesions^6^. This is attributable to low contrast within cortical lesions compared to surrounding normal appearing gray matter (NAGM), normal appearing white matter, and cerebrospinal fluid (CSF)^7^.

7-tesla (T) MRI is the criterion standard in detecting cortical lesions, having demonstrated both better signal-to-noise ratios (SNR) and higher spatial resolution^8^ than 3T imaging. However, cost associated with 7T scanners has limited their availability for clinical practice. Techniques for cortical lesion detection at 3T have relied on advanced imaging methods, such as Double Inversion Recovery (DIR), Phase-Sensitive Inversion Recovery (PSIR), and Inversion Recovery Susceptibility Weighted Imaging with Enhanced T_2_ weighting (IR-SWIET)^6-7,9^. Recently, IR-SWIET demonstrated the highest sensitivity for subpial lesions when compared to other propsed 3T imaging methods^9^. However, these sequences are not readily available on clinical 3T scanners, and time contraints further limit their feasibility in routine care.

T_1_-weighted and T_2-_weighted images display visible signal changes in cortical lesions, although the changes are often subtle. For this reason, the ratio of the signal intensities of these images, (T_1_/T_2_) images, have been explored to potentially accentuate these differences. This ratio has been shown to be sensitive to differences in myelin content while decreasing intensity bias^10^. While debate continues concerning the specificity of the T_1_/T_2_ ratio for cortical myelin^11-13^, T_1_/T_2_ continues to be of interest due to its potential sensitivity to gray matter. Additionally, T_1_/T_2_ changes have been reported in MS^14^. Most importantly, T_1_/T_2_ can be derived from routinely acquired clinical sequences. In this study, we performed a retrospective analysis of MRIs obtained from 10 adults with MS. Cortical lesions were identified at 7T, and contrast within identified cortical lesions was then assessed on multiple 3T sequences with an aim to evaluate the relative sensitivity of T_1_/T_2_ images (MP2RAGE/FLAIR).

## METHODS

### MRI Data

10 adults with MS who participated in a natural history study conducted at the National Institutes of Health and in whom 7T and 3T images, including IR-SWIET, had been previously acquired, were retrospectively included in the study. Detailed information regarding the MRI sequences obtained has been published previously^9^ and is briefly summarized in **Table 1**. 3T MP2RAGE images were acquired on a Siemens Skyra scanner, 3T T_2_w FLAIR and IR-SWIET images were acquired on a Philips Achieva scanner, and 7T images were acquired on a Siemens Magnetom scanner. 3T and 7T Siemens images were acquired a median of 3 months (range 1–7 months) and median 4 months (range 1–7 months) before the 3T Philips scans, respectively.

**Table 1:**
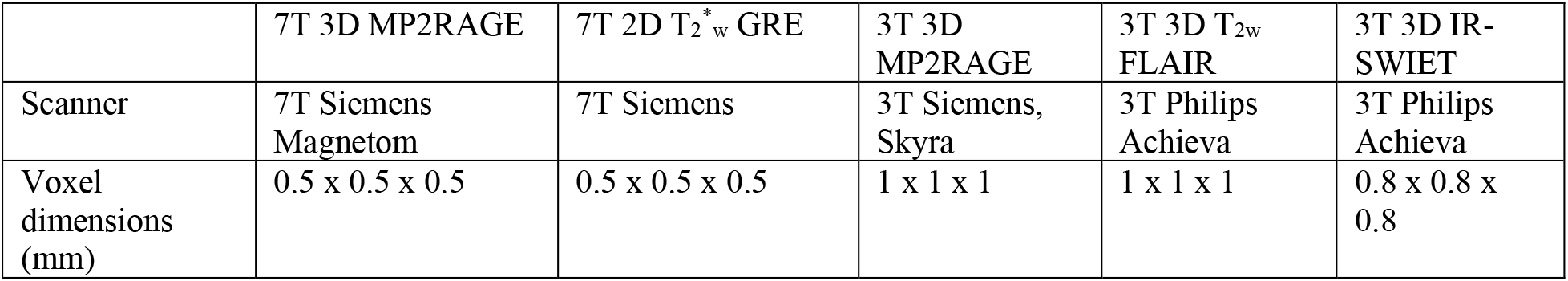
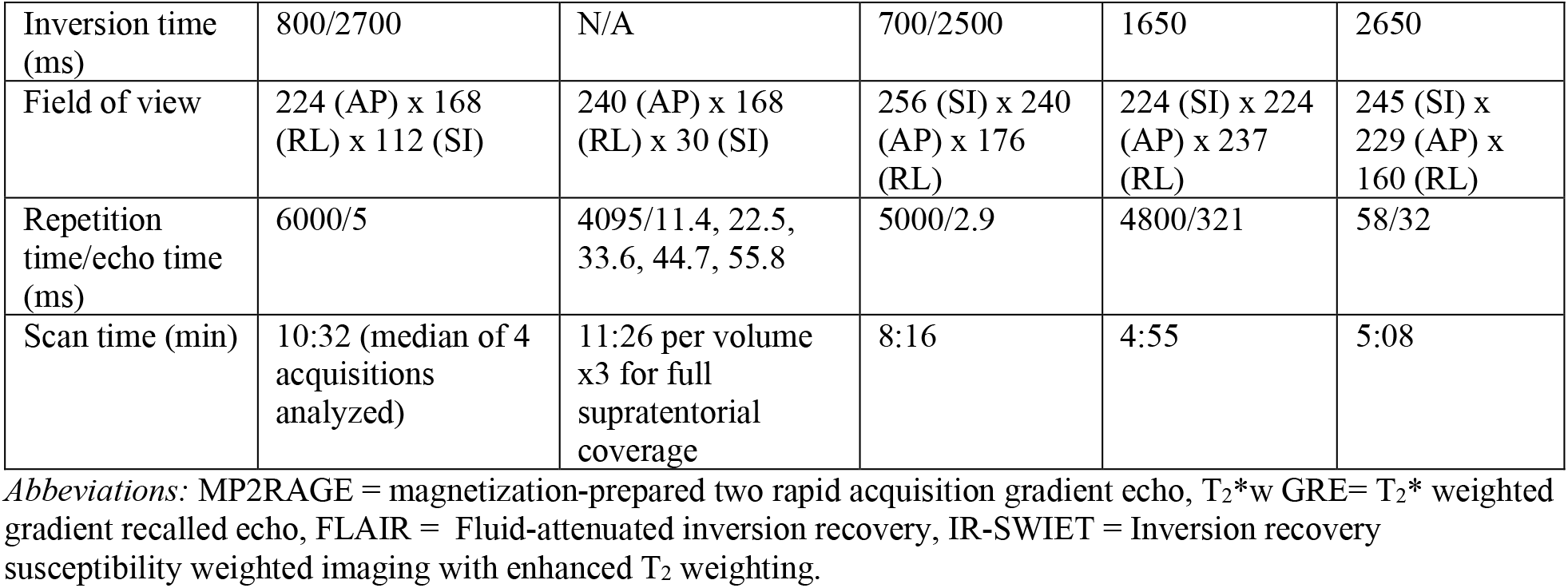
MRI sequence parameters.

### Image Processing

All images as well as previously generated cortical lesion masks were co-registered to the 3T MP2RAGE space, and T_1_/T_2_ images were generated by dividing the 3T T_1_w (uniform denoised) MP2RAGE image by the 3T T_2_w FLAIR image for each subject.

### Image Analysis

Cortical lesion manual segmentations and lesion subtype classification were performed previously using 7T MP2RAGE and T_2_*w GRE images^9^. Cortical lesion subtypes, identified on 7T images as seen in **Figure 1**, were defined as follows: (1) leukocortical: extending across both WM and GM but not touching the pial surface; (2) intracortical: within the cortex without extension to the pial surface or subcortical white matter; and (3) subpial: touching the pial surface of the cortex, with or without WM involvement (**Figure 1**). We included visualization of these lesion subtypes on T_2_/T_1_ ratio images, generated by dividing 3T T_2_w FLAIR images by MP2RAGE images, for additional demonstration of the various cortical lesion subtypes. These images were not included in our analysis, however we would expect results to mirror those of the T_1_/T_2_ ratio images. For each participant, median voxel intensity for each cortical lesion was calculated for each 3T sequence and for the T_1_/T_2_ image.

**Figure 1:**
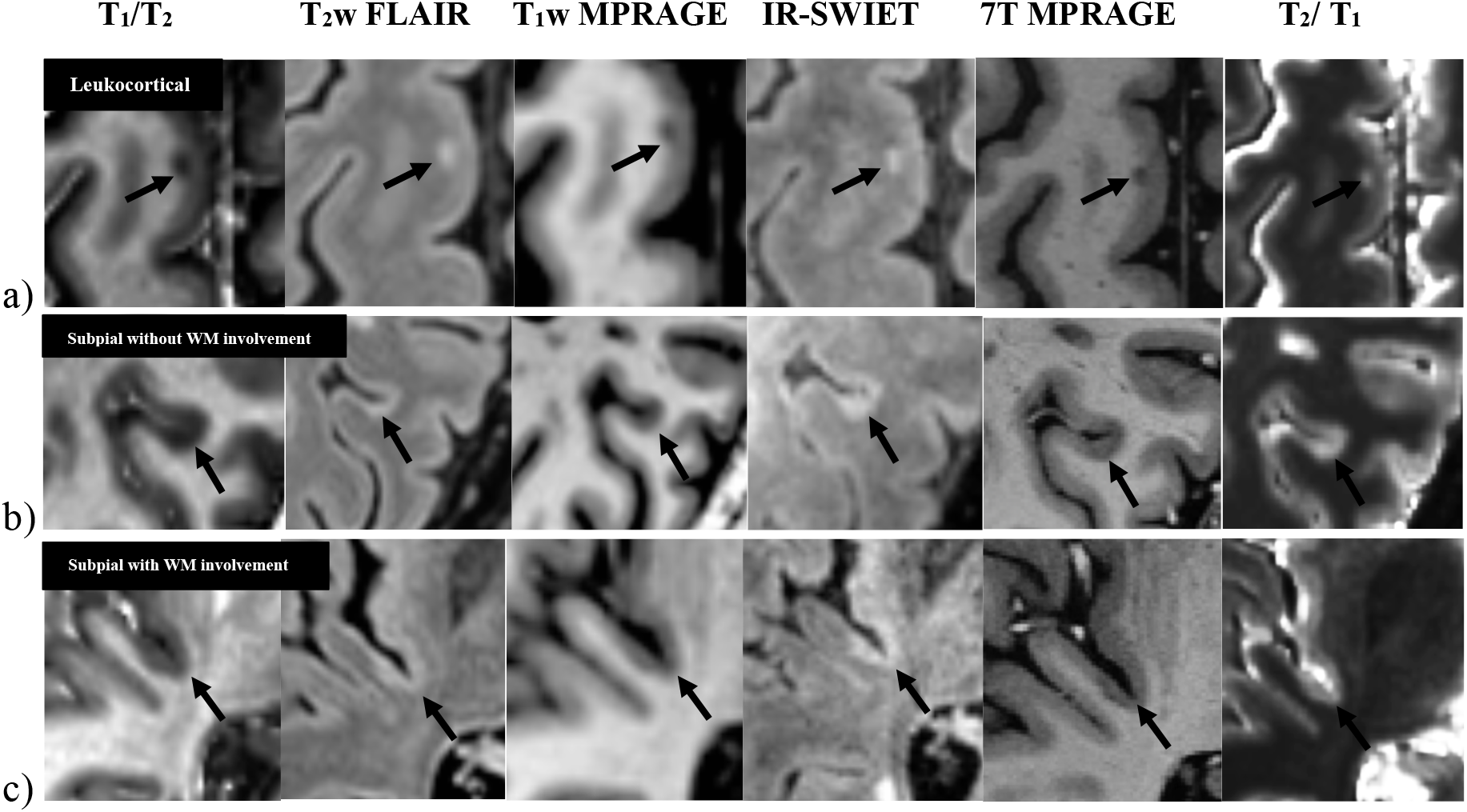
Identification of cortical lesions across modalities. Visualization of (a) leukocortical, (b) subpial without WM involvement, and (c) subpial with WM involvement lesions using T_1_/T_2_, 3T T_2_w FLAIR, 3T T_1_w MP2RAGE, 3T IR-SWIET, 7T MPRAGE, and T_2_/T_1_ images

Lesion segmentations were also used to generate images with lesion-containing voxels removed, to allow for comparisons between lesional tissue and nonlesional normal-appearing tissue. Automated brain segmentations containing 100 identified cortical regions were calculated with 7T MP2RAGE images using the Spatially Localized Atlas Network Tiles (SLANT) algorithm^9,15^. In the coregistered space, these segmentation images were used to determine the cortical region that contained each lesion. Any lesions present in this region were removed using the provided manual segmentations, and the median intensity of the nonlesional region was determined. For each lesion, the given region used to calculate the median nonlesional value was that in which the individual lesion resided. These values were then used to calculate the ratio of median nonlesional cortical signal intensity to median cortical lesion intensity from 3T MP2RAGE and T_1_/T_2_ images, as cortical lesions are hypointense on these images, and the ratio of median cortical lesion intensity to median nonlesional cortical signal intensity for IR-SWIET and FLAIR, as cortical lesions are hyperintense on these images. This ratio is referred to as “cortical lesion contrast.”

### Statistical Analysis

All statistical analyses were conducted using two-sided hypothesis testing and assuming a 5% type I error rate. For each participant, the average cortical lesion contrast was calculated for all lesions and for each subtype of cortical lesion. Paired t-tests were used to test for differences between 3T imaging sequences (MP2RAGE, IR-SWIET, FLAIR, and T_1_/T_2_ image).

### Data Availability

Upon publication and for one year thereafter, deidentified participant level data, including MR images, may be made available upon reasonable request.

## RESULTS

### Study Participants

Images from ten adults (7 women, 3 men) with MS (8 relapsing remitting, 1 secondary progressive, and 1 primary progressive) participated in the study. Mean ± SD age was 48 ± 6 years. Mean ± SD disease duration was 9 ± 5 years. The median expanded disability status score (EDSS) was 2, with a range of 0 to 6.5.

### Cortical Lesion Identification

A total of 614 lesions were identified on 7T T_2_*w and MP2RAGE images (median 47.5 lesions per participant, range 21–128). Per participant median leukocortical lesion number was 17.5 (range 0–42), intracortical lesion number was 2 (range 0–9), and subpial lesion number was 33 (range 9–159). With respect to subpial lesions, the median number with WM involvement was 3 (range 0–59) and without WM involvement 22.5 (range 7–124).

### Quantitative Results

For all cortical lesions identified on 7T images, 3T T_1_/T_2_ demonstrated a larger ratio of median nonlesional cortical signal intensity to median cortical lesion intensity, or “cortical lesion contrast” (median ratio = 1.29, range 1.19 – 1.38), when compared to 3T T_1_w (1.01, 0.97 – 1.10, p<0.002), 3T T_2_w (1.17, 1.07 – 1.26, p<0.002), and 3T IR-SWIET (1.21, 1.01 – 1.29, p<0.03). IR-SWIET demonstrated a greater cortical lesion contrast for all cortical lesions when compared to T_1_w (p<0.01), but did not show a significant difference when compared to T_2_w (p<0.2). Cortical lesion contrast values broken down by individual lesion subtype are shown in **Table 2**.

**Table 2:** Cortical lesion contrast values across modalities. Comparison of cortical lesion contrast values, as defined as the ratio of median nonlesional cortical signal intensity to median cortical lesion intensity for 3T MP2RAGE and T_1_/T_2_ images, as cortical lesions are hypointense on these images, and the ratio of median cortical lesion intensity to median nonlesional cortical signal intensity for IR-SWIET and FLAIR, as cortical lesions are hyperintense on these images. P-values reflect performance of each 3T modality as compared to T_1_/T_2_.

|  | **$T_1/T_2$** |  | **IR-SWIET** |  | **$T_2w$ FLAIR** |  | **$T_1w$ MP2RAGE** |  |
| --- | --- | --- | --- | --- | --- | --- | --- | --- |
|  | Cortical Lesion Contrast | Range | Cortical Lesion Contrast | Range | Cortical Lesion Contrast | Range | Cortical Lesion Contrast | Range |
| All cortical lesions (n = 614) | 1.29 | 1.19 – 1.38) | 1.21 (p<0.03) | 1.01 – 1.29 | 1.17 (p<0.002) | 1.07 – 1.26 | 1.01 (p<0.002) | 0.97 – 1.10 |
| Leukocortical (n = 188) | 1.33 | 1.23 – 1.59 | 1.32 (p<0.04) | 1.13 – 1.35 | 1.25 (p<0.004) | 1.13 – 1.27 | 0.98 (p<0.004) | 0.93 – 1.15 |
| Intracortical (n = 33) | 1.11 | 1.00 – 1.27 | 1.18 (p<0.6) | 0.65 – 1.45 | 1.13 (p<0.7) | 1.08 – 1.22 | 0.90 (p<0.004) | 0.79 – 1.08 |
| Subpial lesions without WM involvement (n = 419) | 1.28 | 1.18 – 1.35 | 1.15 (p<0.01) | 0.97 – 1.27 | 1.15 (p<0.004) | 1.09 – 1.21 | 1.03 (p<0.002) | 0.92 – 1.11 |
| Subpial lesions with WM involvement (n = 124) | 1.24 | 1.12 – 1.40 | 1.18 (p<0.7) | 1.08 – 1.36 | 1.18 (p<0.07) | 1.13 – 1.25 | 0.96 (p<0.04) | 0.91 – 1.19 |

For leukocortical lesions (**Figure 2a)**, T_1_/T_2_ displayed greater cortical lesion contrast when compared to T_2_w (p<0.004), T_1_w (p <0.004), and IR-SWIET images (p<0.04), whereas IR-SWIET was more sensitive when compared to T_1_w (p<0.004) and T_2_w (p<0.02). For intracortical lesions (**Figure 2b**), while T_1_/T_2_ and IR-SWIET continued to display a greater cortical lesion contrast when compared to T_1_w (p<0.004 and p<0.04, respectively), there was no significant difference between T_1_/T_2_ and T_2_w (p=0.65) or T_1_/T_2_ and IR-SWIET (p=0.57).

**Figure 2:**
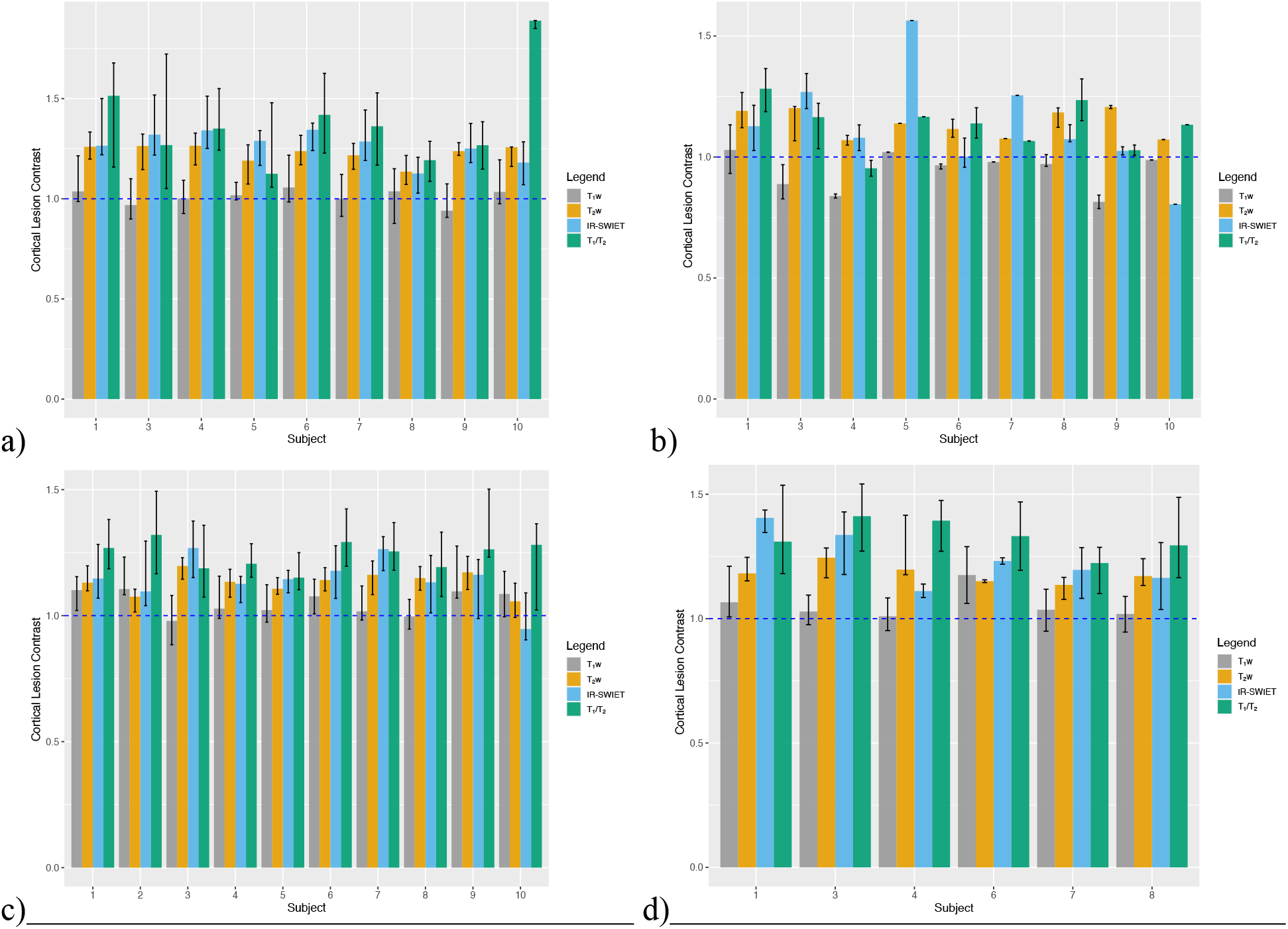
Cortical lesion contrast values compared across modalities. Comparison of identification of (a) leukocortical lesions (n = 188), (b) intracortical lesions (n = 33), (c) subpial lesions without WM involvement (n = 419), and (d) subpial lesions with WM involvement (n = 124) between T_1_w MP2RAGE, T_2_w FLAIR, IR-SWIET, and T_1_/T_2_ across subjects.

When looking at subpial lesions, T_1_/T_2_ displayed greater contrast for lesions that did not extend into the WM (**Figure 2c**) when compared to T_2_w (p<0.004), T_1_w (p<0.002), and IR-SWIET (p<0.01). For subpial lesions with WM involvement (**Figure 2d**), while T_1_/T_2_ continued to display an increase in contrast when compared to T_1_w (p<0.04), there was no significant difference when compared to T_2_w (p=0.06) or IR-SWIET (p=0.69). Additionally, IR-SWIET displayed greater contrast for subpial lesion that did not extend into the WM when compared to T_1_w (p<0.02), however this did not hold true when compared to T_2_w (p=0.92). There was no significant difference for subpial lesions with WM involvement for IR-SWIET when compared to T_2_w (p=0.31) or T_1_w (p=0.06). Specific cortical lesion values for each subtype across modality can be seen in **Table 2**.

## DISCUSSION

In this study, our approach explored the relative contrast of T_1_/T_2_ images (MP2RAGE-derived, T_1_w, uniform denoised image divided by T_2_w-FLAIR) of cortical lesions originally identified on 7T images when compared to 3T T_1_w MP2RAGE, 3T T_2_w FLAIR, and 3T IR-SWIET images. While previous studies exploring the use of T_1_/T_2_ images have utilized 3T T_2_-weighted images for generation of ratio images; we used 3T T_2_w FLAIR images due to potential advantages in increased lesion detection when compared to T_2_-weighted images^16^, although these advantages may be inconsistent across lesion subtype^17,18^. We found T_1_/T_2_ to have greater cortical lesion contrast than that of the individual and T_1_w MP2RAGE sequences for all cortical lesion subtypes. When compared to T_2_w FLAIR and IR-SWIET, T_1_/T_2_ displayed increased contrast for leukocortical lesions and subpial lesions that did not extend ino the WM, however this did not hold true for intracortical lesions and subpial lesions with WM involvement. This suggests that a T_1_/T_2_ image, created from clinical MRI sequences, may be more sensitive to cortical lesions and may be useful for their identification.

Data from previous studies employing T_1_/T_2_ methology and incorporating historipathological correlation have supported its sensitivity for cortical pathological processes in MS^14,20^. There remains some debate concerning whether T_1_/T_2_ may be a better indicator of tissue integrity, rather than a specific marker of cortical myelin status^12-13,19^. While the exploration of T_1_/T_2_ methods has been limited to few studies, these ratio images have been shown to demonstrate sensitivity to cortical neurite density^19,20^ and association with future lesion formation, MS diagnosis, and disability^14^. By contrast, the evaluation of focal cortical damage in MS using T_1_/T_2,_ as pursued in this study, has not been widely evaluated.

Although recently developed 3T imaging methods have improved MS cortical lesion detection, sensitivity for subpial lesions remains a challenge. IR-SWIET has demonstrated increased sensitivity to subpial lesions when compared to other putative approaches to cortical lesion detection, such as other advanced methods like DIR and PSIR^9^. We found T_1_/T_2_ demonstrated increased sensitivity to pre-identified subpial lesions not involving WM when compared to routine 3T methods and IR-SWIET, further suggesting the potential for this approach.

This study had limitations. While the number of participants in this study was small, we analyzed a relatively large number of cortical lesions due to our use of high-resolution, optimized 7T images for their identification. Different MRI scanners were utilized, and the study included scans acquired several months apart. Validation of these findings should include diversity in acquisition parameters in larger cohorts. Future large, prospective studies are needed to verify these results and to evaluate the sensitivity, specificity, and inter-rater reliability of T_1_/T_2_ for MS cortical lesion detection compared to other approaches on 3T.

We found that T_1_/T_2_ was sensitive to cortical MS lesions, and comparable to IR-SWIET, a recently described, sensitive advanced 3T imaging technique for cortical lesion detection. A major strength of the T_1_/T_2_ contrast is that it is calculated from ubiquitously employed, conventional MRI sequences. Our data supports future investigation of the sensitivity of T_1_/T_2_ for cortical pathology in MS.

## Acknowledgements

We thank the NINDS Neuroimmunology Clinic, the NIH Functional Magnetic Resonance Facility, and the NIH Department of Radiology for support with patient care and scanning.

